# A study of the acceptability of a community delirium toolkit to healthcare staff

**DOI:** 10.1101/2025.03.17.25324089

**Authors:** Ashley Gluchowski, Helen Hawley-Hague, Indigo Charles, Helen Pratt, Emma Vardy

## Abstract

**Background:** Delirium is a common condition that frequently results in hospital admission. There is increasing evidence, to show that delirium can be safely assessed and managed in the community, we have previously published the results of a community delirium toolkit pilot demonstrating this.

**Objectives:** We sought to evaluate the acceptability of a community delirium toolkit to healthcare teams involved in its implementation in Greater Manchester (GM). Staff from teams with a high level of adoption were selected.

**Methods:** Eight qualitative structured one-to-one on-line interviews were completed with staff from three teams. Thematic coding was used following a framework analysis.

**Results:** Four themes emerged around set-up (initial contact, support and training), usage (benefits, challenges and embedding in practice), opinion and wider dissemination. Organisational context was key to successful implementation. Staff perceptions of usefulness were an important motivator for staff. New appreciation was acquired around the usefulness of blood tests in investigating causes of suspected delirium. Key enablers were development of a co-produced toolkit and project management with sharing of resources.

**Conclusions:** This study confirms that the GM community delirium toolkit was easy for the teams who engaged with the pilot to use and adopt it.

## Introduction

Delirium is a common condition that frequently results in hospital admission. Delirium is an acute confusion secondary to an underlying physical illness and is associated with significant mortality and economic cost.^1,2^ There is evidence that caring for people in their own home is preferable in terms of prevention^3^ and the suggestion that this may enhance recovery.

Greater Manchester (GM) developed a community delirium toolkit to guide staff in assessment and management of delirium.^4^ The pilot, completed under the Dementia United (DU) programme of work in GM, illustrated the feasibility of an approach assessing and caring for people with delirium in the community with 70% of safely managed at home.

In this study we wished to explore the opinions of healthcare workers in community teams on the implementation and content of the toolkit.

## Methods

### Participant identification

Emails were sent to nurses or Advanced Clinical Practitioners who adopted the delirium toolkit at stages 5,6 or 7 of adoption (see Figure 1 for details of these stages). Higher adopters were chosen as those with most experience of utilising the toolkit. Two staff from each of four pilot areas were approached. Eight health care professionals were interviewed (August-November 2021) across four different areas. Three declined to be interviewed due to time constraints.

### Data collection methods

This was a qualitative project evaluation with structured, one-on-one online interviews conducted by two authors: AG (female, Researcher with a background in Clinical Exercise Physiology) and IC (female, medical student undertaking a research assistant). Neither participants or researcher were known to each other. Interview questions devised by the authors collectively are shown in Table 1. Interviews lasted approximately 30-minutes and were audio recorded using Zoom. No additional notes were taken or repeat interviews carried-out.

**Table 1.**
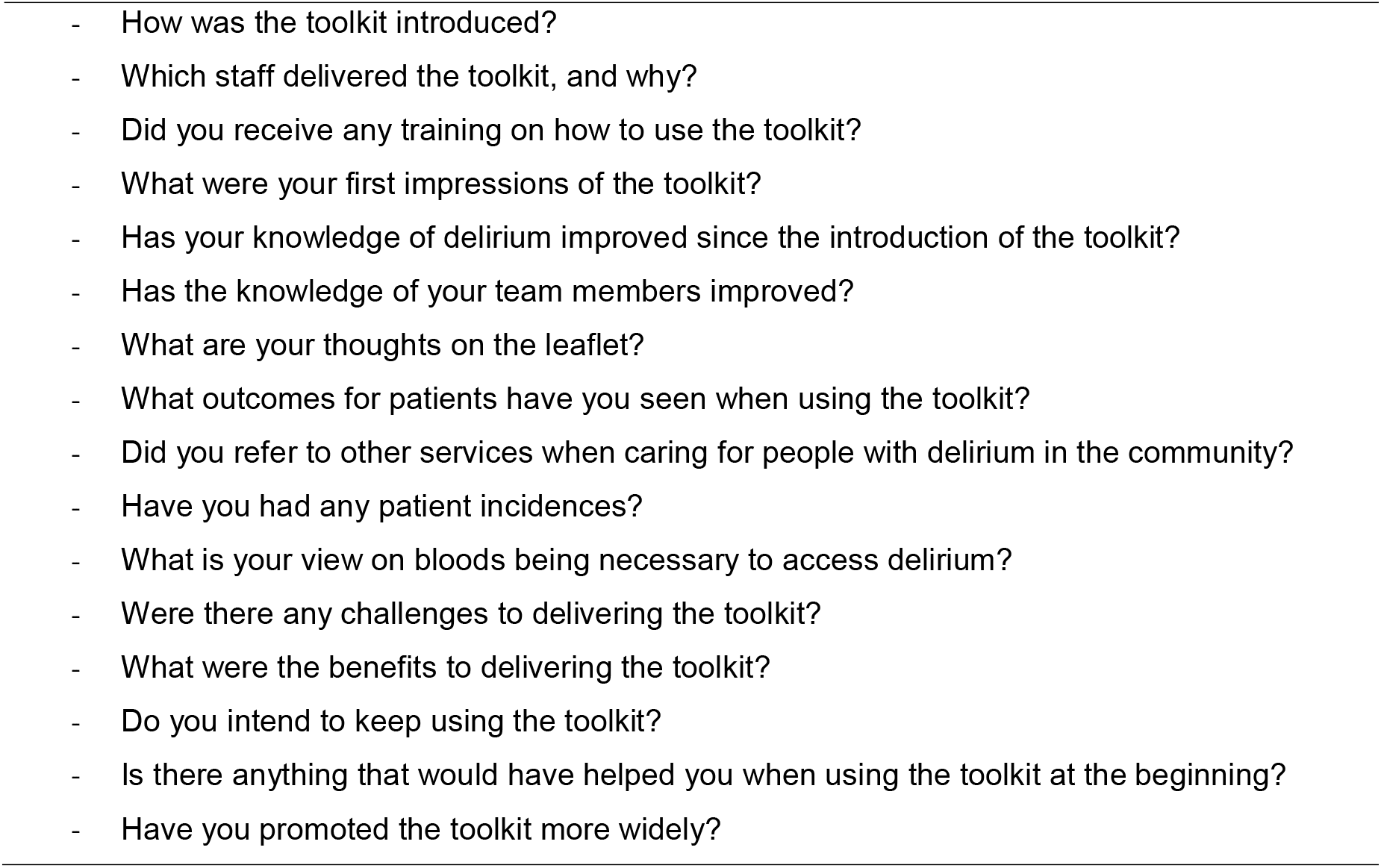
Interview Guide.

The audio recording was transcribed verbatim using a university approved transcription company. Transcripts were not offered for comment or correction.

### Analysis

Thematic coding was used following framework analysis, where the questions around the implementation of the toolkit provided natural structure for the coding.^5^ The NVivo 11 (QSR International) qualitative data analysis software was used to manage the data. Two authors (AG and IC) read and re-read each of the transcripts to immerse themselves further in the data. To increase rigour, a third author (HHH), independently analyzed a random subset of the data to cross-check theme identification, any differences were discussed and consensus reached. This report conforms to the COREQ guidelines (see COREQ checklist).^6^

## Results

From eight interviews, four main themes emerged, summarised in table 2. Quotes are presented to illustrate the findings and identified by site code and number.

**Table 2.** Interview themes.

|  |  |
| --- | --- |
| Delirium community toolkit set-up | Initial contact |
|  | Support |
|  | Training |
| Use of delirium community toolkit | Benefits – accessible resources |
|  | Benefits – knowledge and identification |
|  | Benefits – patients |
|  | Benefits – blood tests |
|  | Benefits – referrals |
|  | Challenges – Right place, right time |
|  | Embedding the toolkit into practice |
| Opinion of toolkit and leaflet | - |
| The wider dissemination of delirium community toolkit | - |

### Theme 1 - Delirium community toolkit set-up

#### Initial contact

DU shared draft versions of the community delirium toolkit and leaflet, as part of the World Delirium Awareness Day event in 2020. From June 2020 onwards expressions of interest to participate in the pilot were received from five locality sites and five different teams. DU offered teams support with launch and feasibility, governance and accountability, and data collection. The sites referenced email as the modality of contact and mentioned their genuine interest in delirium inspired from prior knowledge translation events.

> *I went to the World Delirium Day a couple of years ago now…interest was sought through an email, and one of our advanced practitioners had done delirium as her Master’s*,*… I think, so had in interest. And then myself and another staff member, sort of, felt we were interested in it. So, we had our first meeting, they introduced the toolkit, and we just thought about ways that we could introduce it and we just decided to just do it. (I2)*

#### Support

The teams were provided with an initial launch meeting with support around governance and implementation. This was followed by fortnightly drop-in sessions and sharing of educational resources to support training of the staff and wider team. All meetings occurred virtually due to COVID-19 pandemic restrictions, and were provided by mental health clinician, project manager and consultant geriatrician. Those unable to join sessions were followed up by phone call.

This support provided for the toolkit set-up was discussed as informative, comprehensive, collaborative, and timely.

> *We’ve had meetings really regularly, she’s always really quick to respond and give…. feedback. So, I think the support has been great, and I think you do need that in this, sort of, thing because it’s a new project. (I2)*

#### Training

The sites took a ‘train the trainer’ approach, where one or two leading team members trained all of their staff. The method of training was undertaken due to COVID-19. The approach allowed rapid dissemination and training of large diverse teams which fostered inclusivity, alongside challenges for teams with shift workers, such as finding suitable timings to meet.

> *I feel the information I’ve received, I don’t think it could have been done any other way so I do feel that was a good method. (G1)*
>
> *And we rolled out training quite quickly. All the staff had some form of training. (I2)*

### Theme 2 - Use of community toolkit

#### Benefits – Accessible resources

All toolkit resources were made available to teams by email and then uploaded onto the DU website. The toolkit facilitated and enabled adoption within the community, in part due to its clear and direct protocols, designed for all grades/levels of staff.

> *I think it’s a really good guide and I think all the, you know, the slides that we’d been shared have been really useful because we’ve not had to reinvent something because it’s already there*…*So, it was quite easy really to, you know, navigate and use, so it was really good. (G2)*

Although these documents were uploaded and freely available through the DU website, the suggestion was for better labelling for easier and quicker location of the relevant ones for the specific team.

> *There’s a lot of documents, so it’s looking to find which one are appropriate to your team. Probably signpost to…maybe have like a library of the main documents and supporting documents. (G3)*

#### Benefits – Knowledge and identification

Teams acknowledged that the toolkit helped them to recognize delirium cases earlier, decreasing their time spent in subjective decision making.

> *I think it’s helped to focus on what it is, what can cause it… and how we can intervene. Whereas before it probably…the knowledge was there, but it wasn’t as focused if you like as it is now (F1)*

The toolkit also consolidated language and improved confidence when discussing delirium in the community.

> *the improvement is that we’re able to call it delirium, and when we’re writing letters to GPs, we’ll put delirium secondary to [urinary tract infection], chest infection. We’re recognising if somebody refers in saying off legs, it’s delirium, we’re using the word delirium. (I1)*

#### Benefits – Patients

As a result of increased knowledge and awareness, a delirium diagnosis was considered for patients earlier than previously. An earlier intervention, in turn, has increased the consistency and quality of patient care.

> *I think we are recognising delirium quicker and therefore we’re able to make changes to try and reverse it a little bit quicker. (F1)*

The focus on diagnosis also allowed teams to quickly recognise where other issues were present as well and act more quickly.

> *So not using the (term) delirium however we have recognised patients that are possibly triggering for a little bit of infection and we’ve then gone down the crisis route. Which, although not on the delirium pathway, I suppose left undetected, we could have gone down that route, so I feel it’s been positive in ways that we’ve not been able to quantify as yet*.*(G1)*
>
> *We’ve had one very positive patient experience where we avoided ED attendance and he was able to remain at home with support*.*(G3)*

Moreover, the benefit of the toolkit expanded beyond the individual patient. An increased knowledge and awareness of delirium (and its signs and symptoms) coupled with an earlier diagnosis provided reassurance for families.

> *…it can be quite distressing for the relatives to see their loved ones like that. I think they think, oh, my God, is this it now, you know, is it…? So it’s giving them the information and what to look out for because obviously if they’ve had it once, they’re more likely to get it again and it can take a long time to resolve so not to be afraid of that*..*(I3)*

#### Benefits – Blood tests

With their improved knowledge of delirium healthcare professionals acknowledged their newfound appreciation of the importance of taking investigational blood tests in suspected delirium cases. Staff were able to make informed requests and have the confidence to look for specific markers. Blood draw was now seen as ‘essential.’

> *Before, we might have gone to the GP and said, should we do some bloods, whereas now, because we’ve got it, like, in a process, we can say, we’re going to be doing these bloods because we’ve eliminated everything else, and this is the need. (I1)*

#### Benefits - Referrals

Sites mentioned that referral pathways were more efficient because of earlier awareness and diagnosis.

> *So, what we’ve done is, the district nurses, if they find a delirium we refer them onto Crisis Response Team (CRT) and they will visit literally within hours…CRT will then get involved and either discharge them or stay involved… until the delirium subsides (G2)*

Participation in the toolkit pilot also improved healthcare professionals’ understanding of the scope of other services, thereby enhancing interdisciplinary relationships.

> *I suppose what it has done for our team, because we’ve been working with our crisis response team on this project it’s built that relationship there and given us a little bit more of an understanding of their service and linking into their service. (G1)*

#### Challenges – Right place, right time

For teams, there was discussion surrounding the unique challenge of having one-off interactions with their patients; and for some teams visiting on a set day once a week. This is an important issue given fluctuation of symptoms in delirium.

> *it’s about being in the right place at the right time, certainly for us as district nurses because we might have patients that we only go in to once a week… So it is about capturing them at that right time… the general consensus is it’s easy to follow pathway and shouldn’t present with many problems (G1)*

#### Embedding the toolkit into practice

Subjectively, teams described the toolkit as a success. All teams were able to confidently and definitively say that they intended to continue to use the toolkit. Some teams planned to perform an audit to have more insight on how exactly the toolkit was having an impact.

> *So, you know, hopefully that audit, it won’t show, like I say, massive numbers but it will give us a bit of an idea of what the delirium toolkit has done and what the outcomes of patients were*.*(G2)*

### Theme 3 – Overall opinion of toolkit and leaflet

As part of the community delirium toolkit a patient information leaflet was co-designed and co-produced alongside people with dementia and carers^7^. There were only positive responses by the teams (and throughout their referral networks) around adoption of the toolkit.

> *Everybody was on board, accepting. We’ve not had any feedback from GPs who felt, like, we shouldn’t be doing the bloods or anything. So, I can honestly say, I’ve not had any negative feedback. (I1)*

Additionally, teams appreciated that the toolkit took the evidence around delirium assessment and management and turned it into an easy to remember, simple to use, practical tool.

> *… Obviously delirium’s caused by something and it was just trying to go through that process of trying to eliminate what was the cause which was really good. They had an acronym of PINCH ME. So obviously that was a really useful tool for us to go through that because it was just to cross things off so it was really useful*.*(I3)*

The leaflet developed in tandem with the toolkit^7^, also had positive opinions overall.

> *the feedback’s been really good from the patients that I’ve heard, and the staff have said…it’s the only information that they’ve ever been given about delirium. (I2)*

One team thought that the leaflet was too wordy and suggested making it more concise. The leaflet also presented a challenge to community teams, often working out of the back of their car.

> *…*.*it’s always useful to have leaflets. I think part of the problem is with leaflets is because we’re out and about in the community, we don’t…if you had a stock of all the leaflets you needed, you’d be filling your car boot up. (F1)*

### Theme 4 – Wider dissemination of the toolkit

All teams attempted to disseminate the toolkit to their wider networks. In some cases, specifically to their quality boards,

> *…*.*we’ve done a positive patient experience with the patient, we averted the ED attendance. So that’s been shared at the quality board and also shared it with [name] regarding Greater ManchesterGM*, *and it went to the patient experience group…just again to showcase the good work*.*(G3)*

In other cases, to any service that they thought that might also benefit from adopting the toolkit.

> *…*..*we’ve accessed the OTs and physio just obviously because they see patients and maybe it’s something for them to look out for to refer into our service. We are trying to make better relationships with the GPs as well just so that they know they can refer into our service. If they are worrying that a patient’s got delirium, they don’t necessarily have to send them into hospital*.*(I3)*

Another team suggested it would be beneficial to roll out the toolkit to care homes, since it was perceived that all grades/levels would be able to correctly use the toolkit. If delirium was found, care home staff could refer on to a team to explore causes.

> *I think it is a very useful toolkit that potentially could be rolled out to care homes who are there all the time*.*(G1)*

## Discussion

This qualitative study has for the first time explored the implementation of a delirium toolkit across services within GM in the UK. It provides new evidence of acceptability of the toolkit when used in practice and supports existing evidence from a proof of concept pilot study.^4^ The organisational context, including the training and support provided to support its adoption was key to its success and has been evidenced in previous studies.^8^ Staff perceptions of usefulness within practice was an important motivator for the continued implementation. They felt it increased their knowledge, helped identification of delirium and also enabled them to provide better support and outcomes for patients and their carers. Staff provided suggestions of some changes and additions to the toolkit, which may aid wider spread implementation.

Allowing teams the flexibility to adapt and refine interventions can improve the implementation process, and enhance adoption.^9^ One area where views changed significantly because of the toolkit was around the practice of taking blood samples to investigate causes of delirium.

Initially teams were unclear of the value, but this changed after pilot completion. Language around delirium was consolidated, which was an unexpected benefit of the toolkit.

The implementation of the toolkit could be understood in the context of implementation theory such as the Consolidated Framework for Implementation Research which provides constructs that have been associated with successful implementation.^8^ Our key constructs are: organisational contextual features related to the adaptability construct (the degree to which the toolkit could be refined to meet local need e.g. reorganisation of resources), the champions construct (the teams who supported the role out of the toolkit and implementation), evaluation construct (ability to demonstrate outcomes), and training construct (support for training and train the trainer approach).^10^

Limitations to the study were that sites that did not adopt the community delirium toolkit were not interviewed and may have had less positive reflections or identified other barriers. We were also not able to interview everyone who took part due to resources and individuals capacity to take part and therefore did not meet full data saturation.

Secondly, the pilot study on which this evaluation is based only provides a local and small-scale picture of adoption, influenced by the context of the COVID 19 pandemic

In summary this study confirms that the GM community delirium toolkit was easy for the teams who engaged with the pilot to use and adopt it. Key enablers were development of a co-produced toolkit and project management with sharing of resources. As community services develop, including hospital at home and virtual wards, these findings may be helpful for other clinical teams in implementing delirium care pathways.

## Data Availability

All data produced in the present study are available upon reasonable request to the authors

## Acknowledgements

The community delirium toolkit was co-produced with stakeholders from across health and social care and carers of people with dementia.

With thanks and acknowledgement to the Academic Health Science National Network for the Stages of adoption framework.

Dementia United would like to thank all those people who worked with us over the last four years in supporting the development of the delirium programme of work, which included:

- Delirium task and finish group members including people with lived experience and carers.
- Delegates who attended the annual World Delirium Awareness Day events in 2018, 2019 and 2020; thank you to Michelle Davies Strategic Clinical Networks for her support with these events.
- Dementia Carers Expert Reference Group who very kindly provided initial feedback on the delirium leaflet and the wider engagement of carers in the programme of work.
- With special thanks to NHS Ayrshire and Arran for their community pathway, Healthcare Improvement/Scottish Delirium Association for delirium toolkit and TIME bundle.
- The community teams who engaged with the pilot; Trafford community enhanced care service, Bolton admission avoidance team, Salford urgent care team, Manchester intermediate care service (Gorton South), Stockport Proactive Victoria primary care
- network and wider engaging colleagues such as Salford intermediate care service, North West Ambulance Service.
- Dementia United delirium team Emma May Smith, Lyndsey Kavanagh

## Author contributions

Author contributions: EV and HHH devised the concept of the project. AG and IG conducted the interviews. HHH acted as a third reviewer. AG wrote the methods and results section. HP created the heat map. EV and HP were involved in project pilot implementation. EV and HHH wrote the discussion. AG completed the COREQ form. All authors approved the final paper for submission.

## Ethics statement

### Approvals

The study protocol was approved by local hospital trust research and development teams and deemed to not require ethical approval. We worked with local governance teams to ensure relevant data protection and confidentiality were followed.

## Funding sources

This research is supported by the National Institute for Health Research (NIHR) Applied Research Collaboration Greater Manchester (NIHR200174). The views expressed are those of the authors and not necessarily those of the NHS, the NIHR, the Department of Health and Social Care, or its partner organisations.

## Declaration of Interests

There are no conflicts of interest to declare

## Appendix 1 Stages of adoption heatmap

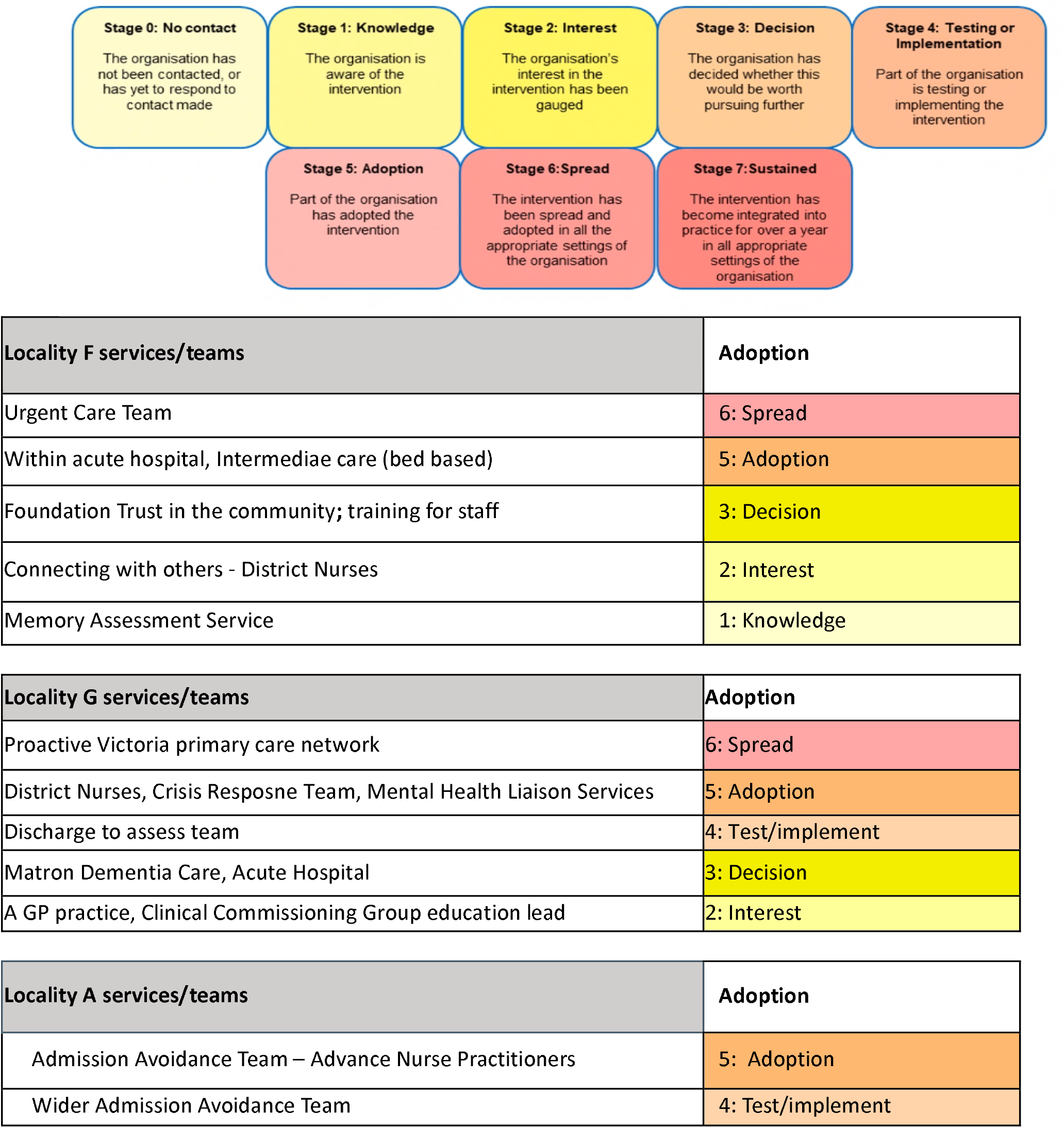
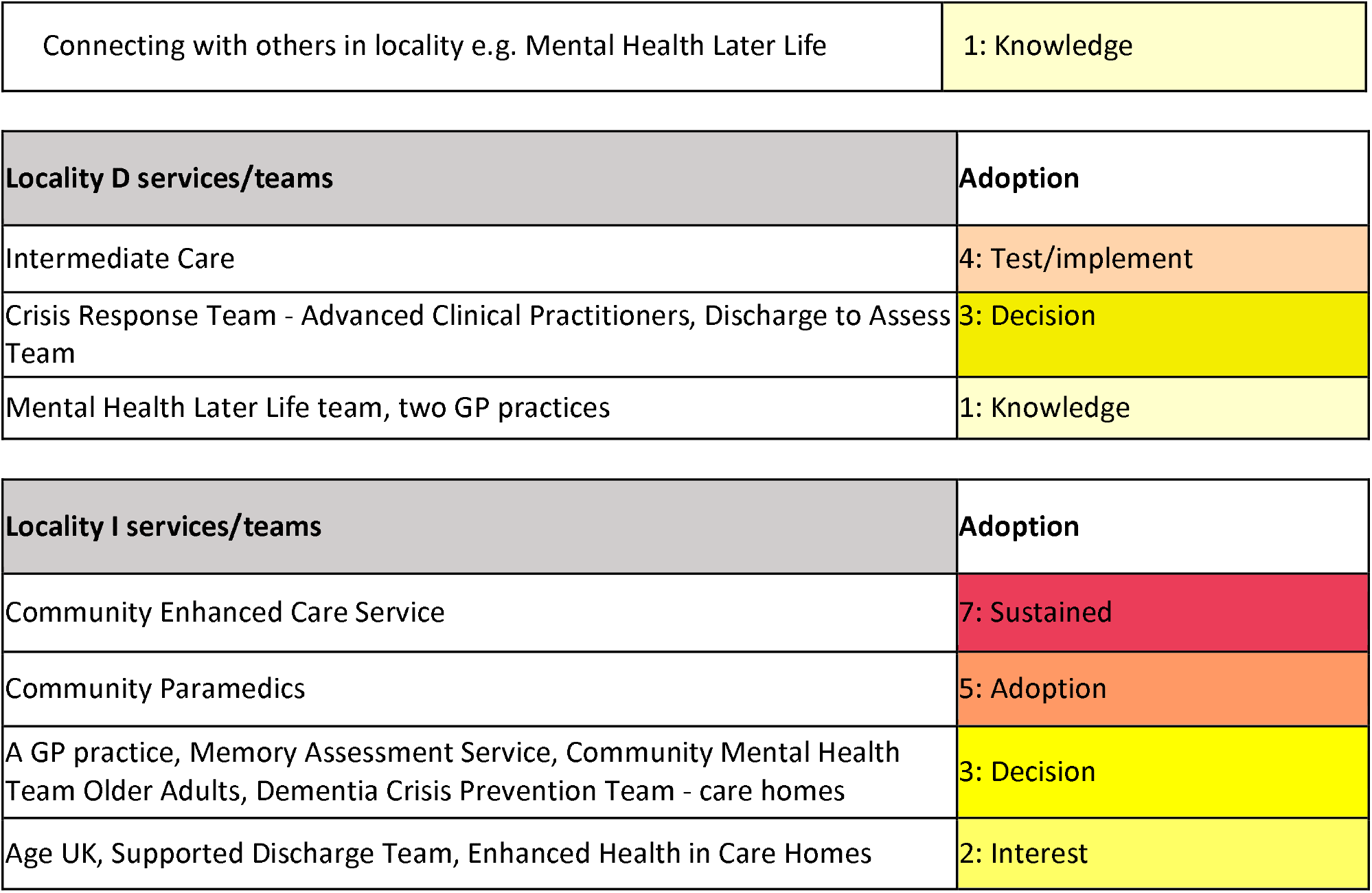

## Notes

### Competing Interest Statement

The authors have declared no competing interest.

## References

1. Scottish Intercollegiate Guidelines Network (SIGN). Risk Reduction and Management of Delirium. 2019. Available from: https://www.sign.ac.uk/our-guidelines/risk-reduction-and-management-of-delirium/ [Accessed 27th October 2022]

2. Caplain GA, Teodorkzuk A, Streatfeild J, Agar MA. The financial and social costs of delirium. Eur Geritatr Med 2019; 11(1): 105–112. DOI: 10.1007/s41999-019-00257-2

3. Shepperd S, Cradduck-Bamford A, Butler C, et al. Hospital at Home admission avoidance with comprehensive geriatric assessment to maintain living at home for people aged 65 years and over: a RCT. Southampton (UK): NIHR Journals Library; January 2022. DOI: 10.3310/HTAF1569

4. Vardy E, Roberts S, Pratt H. Delirium can be safely managed in the community through implementation of a community toolkit: a proof-of-concept pilot study. Future Healthcare Journal 2022; 9(1):83. DOI: 10.7861/fhj.2021-0157

5. Gale NK, Heath G, Cameron E, Rashid S, Redwood S. Using the framework method for the analysis of qualitative data in multi-disciplinary health research. BMC Medical Research Methodology 2013; 13(1):117. DOI: 10.1186/1471-2288-13-117.

6. Tong A, Sainsbury P, Craig J. Consolidated criteria for reporting qualitative research (COREQ): a 32-item checklist for interviews and focus groups. Int J Qual Health Care 2007;19 (6):349–57. DOI: 10.1093/intqhc/mzm042

7. Delirium toolkit leaflet. Dementia United. Available from: https://dementia-united.org.uk/delirium-toolkit-training-resources/ [Accessed 27th October 2022]

8. Li S-A, Jeffs L, Barwick M, Stevens B. Organizational contextual features that influence the implementation of evidence-based practices across healthcare settings: a systematic integrative review. Systematic Reviews. 2018; 7(1):72. DOI: 10.1186/s13643-018-0734-5.

9. Greenhalgh T, Robert G, Macfarlane F, Bate P, Kyriakidou O. Diffusion of innovations in service organizations: systematic review and recommendations. The Milbank Quarterly 2004; 82(4):581–629. DOI: 10.1111/j.0887-378X.2004.00325.x.

10. Damschroder LJ, Aron DC, Keith RE, Kirsh SR, Alexander JA, Lowery JC. Fostering implementation of health services research findings into practice: a consolidated framework for advancing implementation science. Implement Sci 2009; 4(1):50. DOI: 10.1186/1748-5908-4-50.

